# Moral injury and wellbeing in essential workers during the Covid-19 pandemic: Local survey findings

**DOI:** 10.1101/2021.06.14.21257728

**Authors:** Catherine Guy, Edward Kunonga, Angela Kennedy, Paras Patel

## Abstract

Essential workers have faced many difficult situations working during in the pandemic. Staff may feel that they, or other people, have acted wrongly and be distressed by this. This represents moral injury, which has been linked with significant mental ill health.

This survey asked essential workers in County Durham and Darlington about their experiences during the first wave of the pandemic and anything they felt would help. Wellbeing and moral injury were rated using sliders.

There were 566 responses. A majority of respondents reported feeling troubled by other people’s actions they felt were wrong (60% scored over 40, where 0 is “not at all troubled” and 100 “very troubled”, median score=52.5). Respondents were generally less troubled by their own actions (median score=3). Wellbeing and moral injury scores varied by employment sector (e.g. NHS staff were more troubled by the actions of others than non-NHS staff).

Staff suggestions included regular supervisor check-ins, ensuring kindness from everyone, fair rules and enforcement and improving communication and processes. Respondents offered simple, practical actions that could be taken by leaders at team, organisation, societal and government policy levels to tackle moral injury and the underlying causes of moral injurious environments.

Using these findings to develop a strategy to address moral injury is important, not only for staff wellbeing, but staff retention and continued delivery of vital services in these challenging times. Working together, we can seek to reduce and mitigate “moral injury” the same way we do for other physical “injuries” in the workplace.

## Background

The Covid-19 pandemic has devastated many countries and communities across the world. Many governments took actions to prevent onwards transmission and ensure health systems were not overwhelmed.

Staff designated as essential workers have faced not only the direct impact of the pandemic, and potential increased personal risk, but also moral dilemmas when guidance on ways of working contradicted usual standard practice. This was not restricted to health and care but also included staff in local government, education, transport, fire and rescue, policing, shop workers and other settings. Staff may feel that they, or other people, have acted wrongly during the pandemic and be significantly distressed by this. This has been referred to in the literature as moral injury.

According to Greenberg and colleagues, moral injury is defined as distress that results from actions (or inaction) which violate a person’s moral/ethical code. It may involve negative thoughts about themselves or others (for example, ‘I am a terrible person’ or ‘They don’t care about people’s lives’) as well as intense feelings of shame, guilt, or disgust. (1) It is widely agreed that moral injury is not a mental illness but can lead to mental ill health and has been linked with anxiety, depression and suicide. (2)

Moral injury has been reported in around a quarter of healthcare workers in some settings (3) though moral injury amongst essential workers in the UK, particularly during the Covid-19 pandemic, has not been described fully. Seeking staff experiences and views is, therefore, important so that any interventions can be tailored to need and are more likely to be successful. (4)

This survey sought to better understand essential workers’ experience of moral injury, and what they felt would help with the ultimate aim of implementing effective action at all levels of the system. Whole system action, as opposed to individual-focussed approaches, is important since moral injurious environments are created through an interaction of government policy, organisational approaches, team dynamics and broader societal and social pressures. (5)

## Methods

A local multiagency project group was established to address moral injury in essential workers. Members included Tees, Esk and Wear Valley NHS Foundation Trust, County Durham and Darlington NHS Foundation Trust, Durham County Council and Public Health England.

As part of a Needs Assessment and Service Evaluation the group developed a survey to understand moral injury amongst essential workers during the first wave of the Covid-19 pandemic. The group agreed to carry out the survey across County Durham and Darlington and to focus on essential workers defined by government as “critical to the COVID-19 response”. (6)

The survey involved self-administered questions which asked staff about their experiences during the first wave of the pandemic and what they feel would help address these issues. Respondents also used sliders to rate their wellbeing and degree of moral injury they experienced.

The link to the survey (7) was disseminated to staff across all sectors of essential workers by members of the multiagency group and responses collected between 24th September 2020 to 12th October 2020.

### Quantitative analysis

Median scores for moral injury and wellbeing were calculated. Scores in particular employment sectors were compared with scores outside that sector using the Mann-Whitney test. Association between moral injury scores and wellbeing were explored using the Pearson Correlation Coefficient.

### Qualitative analysis

Thematic analysis of free text comments was conducted. Initial codes were compared with independent coding of a section of comments by a second researcher (PP). Codes were combined into themes which were repeatedly reviewed and amended to ensure that they covered the comments provided, were clear in meaning and had the right breadth for effective communication and action.

## Results

There were 566 respondents (79% female, median age group 45-49 years). Respondent employment sectors included NHS/health (54%), social care (17%), local/national government (15%), education/childcare (9%) and others (5%). Detailed demographics, self-reported scores by employment sector and correlation data are included within the supplementary tables. The most frequently skipped scoring question was question 1 regarding how troubled respondents were by their own actions (16 respondents, 2.8%).

### Scores for moral injury and wellbeing

A majority of respondents reported feeling troubled by other people’s actions they felt were wrong (60% scored over 40, where 0 is “not at all troubled” and 100 “very troubled”, and the median score was 52.5).

As Figure 1 shows, respondents were generally less troubled by their own actions (median score 3). Respondents troubled by their own actions also tended to be troubled by other people’s actions (r=0.4091, p<0.0001) with all respondents scoring over 60 for the former, scoring 50 or more for the latter.

**Figure 1:**
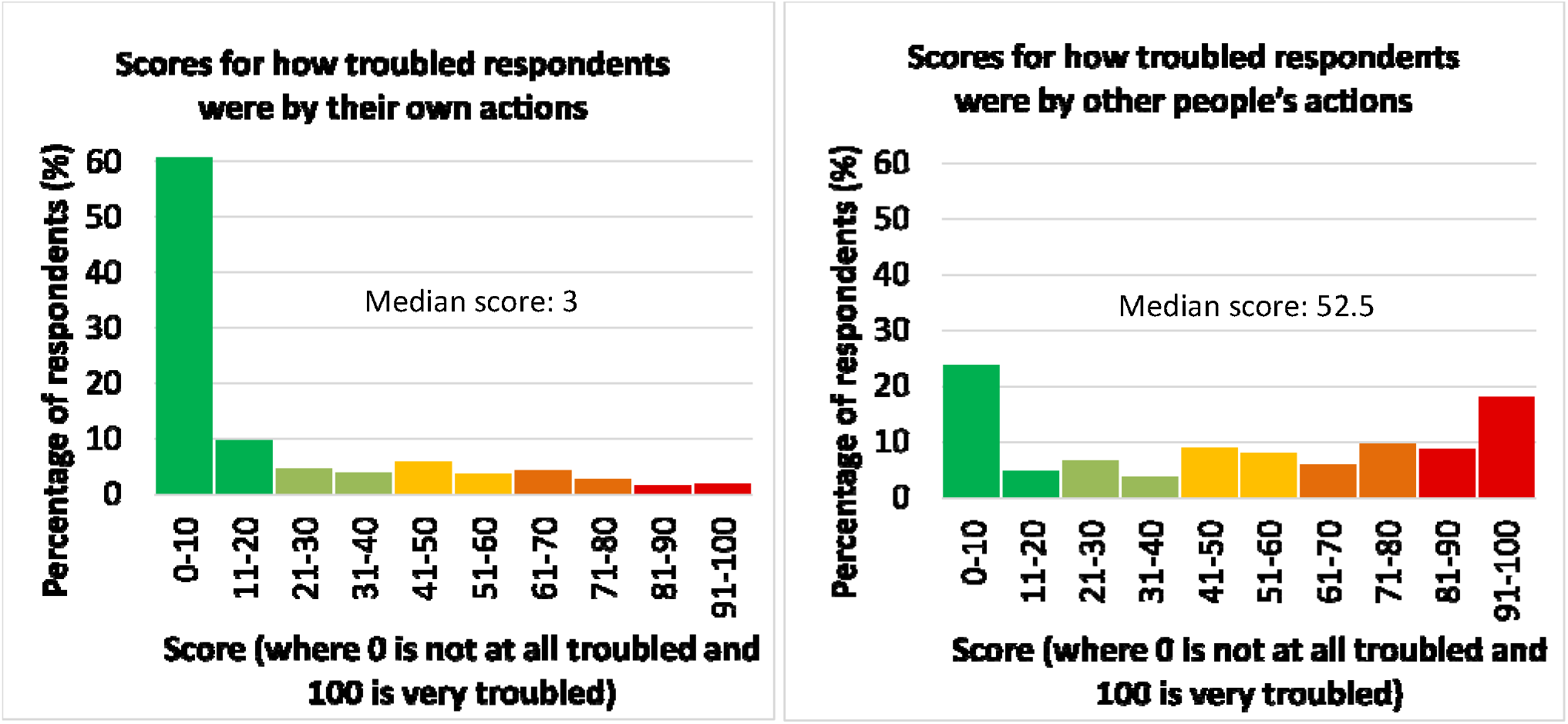
Comparison of how troubled respondents were by their own and other people’s actions.

Both moral injury scores showed statistically significant negative correlations with physical wellbeing (p=0.0008, p<0.0001), mental wellbeing (p<0.0001, p<0.0001) and connectedness with colleagues scores (p=0.0275, p=0.0165).

Overall, median scores for physical and emotional/mental wellbeing were 70 and 52.5 respectively (where 0 is “very poor” and 100 “excellent”). The median scores for connectedness with colleagues and connectedness with others outside of work were 54 and 48 (where 0 is “not close/very isolated” and 100 “very close/connected”).

### Variation in scores by employment sector

Across employment sectors, there were statistically significant differences in moral injury and wellbeing:

- NHS/Health staff reported being more troubled by the actions of others (median score 58) than non-NHS staff (p=0.0334). They also reported lower mental wellbeing (median score 50) than non-NHS staff (p=0.0004).
- Social care sector staff generally reported feeling more connected to colleagues (median score 70) than non-social care staff (p=0.0010).
- Local or national government staff reported higher mental wellbeing (median score 63) than non-government staff (p=0.0472).
- Education/childcare staff reported higher mental wellbeing (median score 69.5, p=0.0082) and higher physical wellbeing (median score 81, p<0.0001) than non-Education/childcare staff.
- Staff in sectors with few respondents (n=31) reported feeling more troubled by other people’s actions (median score 71) compared with staff in NHS/health, social care, government and education/childcare sectors (p=0.0408). This category of staff included key public services, essential goods production or sale and public safety.

### What troubled respondents and staff suggestions

Table 1 summarises actions or situations that troubled staff, and their suggestions, as well as prompts for actions organisational change can take time and the importance of empowering everyone to make a difference was noted. Examples of individual quotes were also shared with local organisations as a powerful way of promoting change. (8)

**Table 1:** Qualitative themes and prompts for action.

| Prevention Theme | What troubled staff | Staff suggestions | Prompts for action |
| --- | --- | --- | --- |
| **Supervisor check-ins** | Fewer opportunities to connect with their manager/supervisor was noted. This created a sense of isolation, lack of value and lack of opportunity to raise issues that could be dealt with. | Regular supportive check ins with managers, informal team meetings and accessible “Mental Health Support” | Who will you contact to let them know what you, or your staff, need from them to function well?<br>How can you connect with staff to meet their needs and concerns? |
| **Kindness** | Receiving “Verbal abuse from colleagues due to their own stress levels”, being “told off” for raising “serious safety concerns” and a “public perception that nurses are carrying disease ” | Educating the public, “Challenging toxic behaviours within teams” and support for senior staff “to manage their own emotions”. It was noted that a “command and control” approach “needs to be balanced with kindness and compassion” | What one small thing can you do, for one other person today, that might help them feel supported and connected?<br>What would help you to stay in enough control of your emotions not to pass your stress on? |
| **Fair rules & enforcement** | Feeling frustrated or angry about rules not being obeyed by colleagues, people in authority and the public. There were differing views about when people should work from home and what fair sharing of workload means. | Rules “need to be really clear, reasonable” and “enforced” with “spot checks of buildings”. Choices, collaboration and ample notice about role allocation. People asked that policies “apply to all” but also that “individual needs” are taken in to account. | Who do you need to have a conversation with to better understand each other’s needs and values?<br>How are you ensuring rules are followed fairly?<br>...and balancing that with individual need? |
| **Effective communication & processes (the way things are done)** | A “lack of communication” from government, organisations, leaders and between agencies or team members. Being worried about the way things are done, patient care and the safety of staff and apparent lack of action on issues raised. | Clearer communication and opportunity to raise concerns and find solutions. Staff had some good specific ideas e.g. have a “designated social worker” for each care home, use of “NHS mail to exchange information quickly” and provision of adequate requested resources e.g. PPE, uniform, equipment. | What can you do to share ideas/solutions/feedback or decisions clearly at the right time with the right people? |

Some themes appeared specific to particular staff groups. For example, concerns about patient, resident or client care were particularly reported by health and social care staff. These concerns included feeling that elderly people were being “written off” and refused care or that “vital info is being missed” by only contacting people by telephone.

## Discussion

This survey met the aim of better understanding essential workers’ experience of moral injury, and what they felt would help. The results suggest moral injury is a significant issue for many essential workers which fits with research of other staff groups. (9) It is interesting, however, that staff reported being more troubled by the actions of other people, than their own actions, since examples involving the latter may be easier to convey sympathetically than disgust or anger. (10)

Variation in moral injury and wellbeing across different employment sectors could suggest areas for further investigation or targeting of interventions. However, distress was present across all sectors and it is unclear how scores compare to the general population.

Some staff suggestions (regular supervisor check-ins, kindness from everyone, better communication and changes to the way things are done) have been mentioned previously in moral injury literature (1) or staff wellbeing guidance. (11) Other suggestions, including focussing on fair rules/enforcement, may reflect the pandemic response.

Since respondents in this survey suggested many practical actions that a variety of stakeholders could take, moral injury should be addressed at all levels of the system. Whilst individual approaches (e.g. support after moral injury occurs) may be seen as the easiest to implement, they should not be at the expense of team, organisation, societal and government policy interventions which tackle the underlying causes of moral injurious environments. (5)

A draft framework for action to reduce and mitigate moral injury is under development locally (8) but a wider strategy to address moral injury is critical, not only for staff wellbeing, but also for staff retention and continuing delivery of vital services in these challenging times. Such a strategy could be made more effective by further developing the evidence base on both ways to reduce moral injurious events, and the best interventions to address moral injury after it happens (including moral injury caused by other people’s actions).

This survey has several limitations including being limited to one area in northern England, variation in response rates across employment sectors and use simple of self-rated sliders for moral injury and wellbeing. However, respondent suggestions may provide ideas for simple actions, which represent good practice, for leaders looking after the wellbeing of staff. Working together, we can seek to reduce and mitigate moral injury in the same way we do for physical injuries inside and outside the workplace.

## Supporting information

Supplemmentary table: moral injury and wellbeing scores by employment sector

Supplementary table: corellation of wellbeing and moral injury scores

Supplementary table: summary of demographics

## Data Availability

Example quotes available at https://www.recoverycollegeonline.co.uk/your-mental-health/coronavirus/for-staff/moral-injury-and-staff-wellbeing/ All survey data stored securely to maintain anonymity

There are no competing interests for any author

## Acknowledgements

Thanks to members of the multiagency group who contributed to the design and dissemination of the survey, including Lyn Williams, Jo Murray, Andrew Moore, Jane Sunter, Clare Leeds, Lucy Treanor and Alison Brabban.

