## Supplementary material for "Moral injury and wellbeing in essential workers during the Covid-19 pandemic: Local survey findings": Supplemmentary table: moral injury and wellbeing scores by employment sector

**Median scores by employment sector**

| Score (0-100) | All sectors (n=564) | NHS/ Health (n=305) | Social Care (n=95) | Local/national Government (n=83) | Education/ Childcare (n=50) | Other sectors (n=31) |
| --- | --- | --- | --- | --- | --- | --- |
| How troubled by own actions | 3 | 4 | 1 | 1 | 1 | 15 |
| How troubled by others actions  (Kruskal-Wallis test across sectors p=0.0090) | 52.5 | 58*  p=0.0334 | 55 | 40 | 30 | 71*  p=0.0408 |
| Physical wellbeing  (Kruskal-Wallis test across sectors p=0.0008) | 70 | 70 | 76 | 67 | 81**  P<0.0001 | 60 |
| Mental wellbeing  (Kruskal-Wallis test across sectors p=0.0003) | 52.5 | 50* p=0.0004 | 59 | 63**  p=0.0472 | 69.5**  p=0.0082 | 50 |
| How connected with colleagues (Kruskal-Wallis test across sectors p=0.00215) | 54 | 50 | 70** p=0.0010 | 50 | 60 | 59 |
| How connected with family/ friends/ others | 48 | 46 | 50 | 42.5 | 50 | 42 |

Higher scores are “worse” for moral injury but lower scores are “worse” for wellbeing/connectedness

Red type and * denotes scores for significantly worse than respondents outside that sector and green or ** significantly better (where p<0.05 using the Mann-Whitney test).

Before comparing individual sectors, the difference in scores across all employment sectors was investigated using the Kruskal-Wallis test (see first column).
