## Supplementary table: corellation of wellbeing and moral injury scores for "Moral injury and wellbeing in essential workers during the Covid-19 pandemic: Local survey findings"

|  | How troubled by own actions score | How troubled by other people’s actions score | Physical wellbeing score | Mental wellbeing score | Connectedness with colleagues score | Connectedness with people outside work score |
| --- | --- | --- | --- | --- | --- | --- |
| How troubled by own actions score | 1 |  |  |  |  |  |
| How troubled by other people’s actions score | 0.4091* (p<0.0001) | 1 |  |  |  |  |
| Physical wellbeing score | -0.1428* (p=0.0008) | -0.2228* (p<0.0001) | 1 |  |  |  |
| Mental wellbeing score | -0.2436* (p<0.0001) | -0.3276* (p<0.0001) | 0.5496* (p<0.0001) | 1 |  |  |
| Connectedness with colleagues score | -0.0947* (p=0.0275) | -0.1022* (p=0.0165) | 0.2496* (p<0.0001) | 0.3359* (p<0.0001) | 1 |  |
| Connectedness with people outside work score | -0.0733 (p=0.0869) | -0.1144* (p=0.0070) | 0.1914* (p<0.0001) | 0.2771* (p<0.0001) | 0.2301* (p<0.0001) | 1 |

Supplementary table: Correlation of moral injury and wellbeing scores
