## Supplementary table: summary of demographics for "Moral injury and wellbeing in essential workers during the Covid-19 pandemic: Local survey findings"

| **Demographics or other characteristics of respondents** | **Percentage of respondents (and number of respondents in brackets) unless otherwise specified** |
| --- | --- |
| **Age:** n=561 median age group (range) | Median 45-49 years (mode 50-54 years, range from “24 or under” to “70 or older”) |
| **Gender:** n=561 |  |
| Female | 79.32% (445) |
| Male | 18.00% (101) |
| Prefer not to say | 2.67% (15) |
| Prefer to self describe | 0 |
| **Ethnicity:** n=563 |  |
| White | 95.20% (536) |
| Don't know or prefer not to say | 2.49% (14) |
| Other ethnic groups (including Mixed, Black, Asian or Other) | 2.30% (13) |
| **Main area of work:** n=562 |  |
| County Durham | 77.22% (434) |
| Darlington | 14.06% (79) |
| Across County Durham and Darlington equally | 6.58% (37) |
| Other area in North East | 2.14% (12) |
| **Do you have any physical or mental health conditions, illnesses, or disabilities lasting or expected to last 12 months or more?:** n=565 |  |
| Yes | 33.10% (187) |
| No | 60.00% (339) |
| Don't know or prefer not to say | 6.90% (39) |
| **Sector of work:** n=564 |  |
| The NHS or health sector | 54.08% (305) |
| Social Care (including care home staff and domiciliary care) | 16.84% (95) |
| Local/national government | 14.72% (83) |
| Education and Childcare | 8.87% (50) |
| Key public services (e.g. justice system, religious staff, frontline charities, staff caring for the deceased, journalists and broadcasters) | 3.19% (18) |
| Other sectors including: Food or other essential goods production/ processing/ distribution/ sale/ delivery, Public safety (e.g. police, fire, prison staff), Transport, Utilities, communication, or financial services or other | 2.30% (13) |
